# A Systematic Review and Meta-analysis of Pregnancy and COVID-19: Signs and Symptoms, Laboratory Tests and Perinatal Outcomes

**DOI:** 10.1101/2020.09.28.20202945

**Authors:** Soheil Hassanipour, Saeed Bagheri Faradonbeh, Khalil Momeni, Zahra Heidarifard, Mohammad-Javad Khosousi, Leila Khosousi, Hosein Ameri, Morteza Arab-Zozani

**Affiliations:** Gastrointestinal and Liver Diseases Research Center, Guilan University of Medical Sciences, Rasht, Iran; Health Management and Economics Research Center, Iran University of Medical Sciences, Tehran, Iran; Department of Public Health, Faculty of Health, Ilam University of Medical Sciences, Ilam, Iran; Msc of HTA, Tehran University of Medical Sciences, Tehran, Iran; Caspian Digestive Disease Research Center, Guilan University of Medical Sciences, Rasht, Iran; Reproductive Health Research Center, Guilan University of Medical Sciences, Rasht, Iran; Health Policy and Management Research Center, School of Public Health, Shahid Sadoughi University of Medical Sciences, Yazd, Iran; Social Determinants of Health Research Center, Birjand University of Medical Sciences, Birjand, Iran

**Keywords:** COVID-19, Pregnancy, Diagnosis, Meta-analysis

## Abstract

**Background:** COVID-19 caused by severe acute respiratory syndrome coronavirus 2 appeared in December 2019 in Wuhan, China.

**Objective:** We aimed to investigate the clinical manifestation include signs and symptoms, laboratory results, and perinatal outcomes in pregnant women with COVID-19.

**Materials and Methods:** We searched PubMed via LitCovid hub, Embase, Scopus, Web of sciences, and Google scholar on 07 April 2020. Meta-analysis was performed via CMA software using the Mantel-Haenszel method. The event rate with 95% CI was calculated for each variable.

**Results:** Ten studies were selected. The pooled prevalence for fever, post-partum fever, cough, myalgia, fatigue, dyspnea, sore throat, and diarrhea were 66.8 %, 37.1 %, 35.5 %, 24.6 %, 14.9%, 14.6 %, 11.5%, and 7.6 %, respectively. Laboratory test results were 49.8 % for lymphopenia, 47.7 % for leukocytosis, 83.7 % for elevated neutrophil ratio, 57 % for elevated C-reactive protein, and 71.4 % for decreased lymphocyte ratio. The rate of cesarean section for delivery in all cases was 84%. Only one case was the newborn of a mother with COVID-19 positive. Also, there was only one death due to Decreased lymphocyte ratio.

**Conclusion:** Fever was the most common signs and symptoms in pregnant women with COVID-19. Among the laboratory tests, the highest amount was related to elevated neutrophil ratio. It seems that due to the differences between pregnant women and the general population, special measures should be considered to treat these patients.

## Introduction

Coronavirus Disease 2019 (COVID-19) caused by Severe acute respiratory syndrome coronavirus 2 (SARS-CoV-2) appeared for the first time in December 2019 in Wuhan, China. The disease then spread rapidly around the world to the point where it was confirmed a pandemic by the World Health Organization (WHO) (1, 2). COVID-19 is an infectious disease with respiratory symptoms almost similar to SARS (2003) and MERS (2012) epidemics (3, 4). In some cases, the disease can lead to a sensitive respiratory condition, many of which require specialized management in the intensive care unit (ICU) (5).

Respiratory droplets along with close contact transmission are the considerable routes of transmission. There is also a possibility of aerosol transmission in a close environment when exposed to high concentrations of aerosol for a protracted period (6). On the other hand, touching surfaces or objects that are touched by an infected person can transmit the disease (7). Studies have also shown that older age and comorbidity play an important role in determining the severity and clinical consequences of the disease (8).

Because most studies have focused on patients infected with the new coronavirus in the general population, bounded details are available regarding pregnancy outcomes in women infected with COVID-19. It has caused particular concern among pregnant women, as both SARS-CoV and MERS-CoV viruses have been shown to cause severe side effects in pregnant women (9, 10). A study by Wong and colleagues examined pregnant women with SARS in 2004 in Hong Kong and found that pregnant women showed high rates of death and mortality (11). Also, a study by Mertz et al has been shown that women infected with influenza were more at risk than healthy pregnant women (12). Chen and co-authors also reported that pregnancy with pneumonia could be associated with the risk of cesarean delivery, preterm delivery, a decrease in the baby’s Apgar score, and weight loss at birth, and so on (13).

It is obvious that parturient has relatively depressed immunity or immune suppression and in theory, it could be more at risk of contracting the virus. Also, confronting SARS-CoV-2 during pregnancy is a serious threat to pregnant women and their fetuses (14, 15). Therefore, during the prevalence of COVID-19, a disease without approved treatment, it is important to prevent pregnant women from becoming infected during the epidemic/pandemic period.

Pregnant women are at risk of infection to respiratory pathogens and severe pneumonia because they are in an immunosuppressive state and changes in physiological adaptation during pregnancy (eg, increased diaphragm levels, increased oxygen consumption) can cause hypoxia intolerance in these patients. For example, the outbreak of influenza in 1918 caused total mortality of 2.6% in the population, but in pregnant women, it was about 37% (16). It has also been stated that pregnant women are at increased risk of complications from the H1N1 epidemic influenza virus infection in 2009 and have been hospitalized more than four times as often as other patients (relative risk 4.3 95% CI: 2.3– 7.8) (17). Therefore, it is important to study COVID-19 signs and symptoms in pregnant women with the disease and its effects on newborns are very important, and therefore this study is aimed at investigating the clinical manifestation include signs and symptoms, laboratory results, and prenatal outcomes in pregnant women with COVID-19.

## Methods

This systematic review and meta-analysis followed by the Preferred Reporting Items for Systematic Reviews and Meta-Analysis (PRISMA) statement (18).

### Eligibility criteria

All studies were included based on the following criteria: they investigated COVID-19 in pregnant women or during pregnancy and were in the English language. Studies were excluded if the researchers haven’t access to the full-text of the article or the data about the outcomes were not sufficient. Also, we exclude studies without the peer review process.

### Information sources and search

We searched PubMed via LitCovid hub, Embase, Scopus, Web of sciences and Google scholar using a specific search strategy (“2019 nCoV” OR 2019nCoV OR “2019 novel coronavirus” OR “COVID 19” OR COVID19 OR “new coronavirus” OR “novel coronavirus” OR “SARS CoV-2” OR (Wuhan AND coronavirus) OR “COVID 19” OR “SARS-CoV” OR “2019-nCoV” OR “SARS-CoV-2” AND pregnancy OR “pregnant women”) on 07 April 2020. Our search was not limited based on items such as type of study or publication date but limited to studies with full-text in the English language. We also search for the references of included studies for capturing potential studies in this field. In terms of incomplete data, we contacted the corresponding author of the article for more information.

### Study selection

After importing the records to EndNote X7, the duplicate records were removed and then screened based on the title, abstract, and full-text considering the eligibility criteria. All stages were conducted using two independent reviewers and the potential disagreements were solved through consultation with a third reviewer.

### Quality appraisal

Two independent reviewers assessed the included studies for quality issues. Because the final studies were case-series and case-control, the JBI checklists related to this type of study were used. These checklists include 10 questions for case-control and case-series studies. These questions investigate issue regarding domain such as inclusion criteria, reliability and validity of methods, sampling process, transparency in data and results, and statistical analysis. The detail about each question mentioned at the end of the questionnaire (Supplementary 1). We score one for yes and zero for no in each question. (19, 20).

### Outcomes measures

The investigated outcomes were signs and symptoms (cough, diarrhea, dyspnea, fatigue, fever, myalgia, sore throat, and post-partum fever), laboratory test results (lymphopenia, leukocytosis, elevated neutrophil ratio, elevated C-reactive protein, and decreased lymphocyte ration), type of delivery (cesarean), and perinatal outcomes (COVID-19 positive, low birth weight, premature, complication, and died). For all outcome variables, we extract the number of events and sample size.

### Data analyses

Meta-analysis was performed for signs and symptoms, laboratory tests, and type of delivery using the event rate (the proportion of the occurrence of an event in the subjects to the total subjects under study) with CMA (Version 2) software using the Mantel-Haenszel method. Also, we used narrative synthesis for reporting the results of the perinatal outcome. The Q-value was applied to discover between-study heterogeneity, and *I*^2^ values were calculated to assess statistical heterogeneity. We used random-effect model based on the level of heterogeneity. In the following of Cochrane criteria if the heterogeneity was ≥50 we used the random-effect model (21). The event rate with 95% CI was calculated for each variable. We used Egger’s test and visual inspection of the funnel plot for assessing publication bias. Also, we conducted a meta-regression for an association between mean age and each sign and symptoms, laboratory test, and type of delivery.

## Results

### Description of search

After searching all international databases, 4721 articles were found and after removing duplicate articles, 3985 articles were examined in terms of title and abstract, out of which 17 articles entered the next stage. After reviewing the full texts of the articles, 10 articles entered the systematic review (22-31). In the screening stages of studies, they were excluded for a variety of reasons, which included unrelated topics (2 articles), unassociated population (4 articles), and duplicated study (1 article). The overall sample size of the included studies was 135 Pregnant women with COVID-19 diagnosis. The flowchart of the included studies is presented in Figure 1.

**Figure 1:**
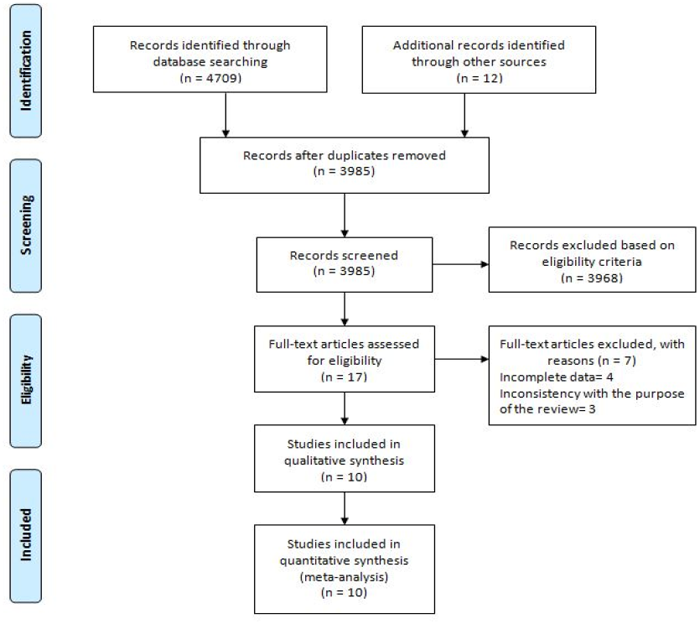
Study selection flow diagram

### Characteristics of included studies

Based on the geographical location, all included studies were performed in China. The summary characteristics of the included studies were shown (Table I).

**Table 1.** Basic information of included studies

| Author, year | Country | Setting | Time period | Study design | Sample size | Mean age (Yr) (Range) | Mean gestational age (Wk) (Range) |
| --- | --- | --- | --- | --- | --- | --- | --- |
| Chen et al, 2020-1 (23) | China | Renmin hospital | From 30 January to 23 February 2020 | Case series | 17 | 29.5 | < 37=3<br>>37= 14 |
| Chen et al, 2020-2 (24) | China | Maternal and Child Hospital of | From January 20 and February 10, | Case series | 5 | 28.4 (25-31) | 38-40 |
|  |  | Hubei Province | 2020 |  |  |  |  |
| Chen et al, 2020-3 (22) | China | Zhongnan Hospital | From Jan 20 to Jan 31, 2020 | Case series | 9 | 32.5 (26-40) | 36-39 |
| Khan et al, 2020 (25) | China | Renmin hospital | From Jan 28 to March 1, 2020 | Case series | 3 | 29.3 (27-33) | (34-39) |
| Li et al, 2020 (26) | China | Hubei Provincial Maternal and Child Health Center | From January 24 to February 29, 2020 | Case-control | 16 | 30.9 | 38 |
| Liu et al, 2020-1 (29) | China | Hospitals outside of Wuhan | From December 8, 2019, to February 25, 2020 | Case series | 13 | 29.6 (22-36) | (25-38) |
| Liu et al, 2020-2 (28) | China | Multi-center | from January 27 to February 14, 2020 | Case-control | 41 | 30.5 (22-42) | NR |
| Liu et al, 2020-3 (27) | China | Union Hospital | from January 20, 2020, to February 10, 2020 | Case series | 15 | 32 (23-40) | 32 (12-38) |
| Yu et al, 2020 (30) | China | Tongji Hospital | From Jan 1 to Feb 8, 2020 | Case series | 7 | 32 (29-34 ) | 39 (37-41) |
| Zhu et al, 2020 (31) | China | Multi-center (5 hospitals) | From January 20 to February 5, 2020 | Case series | 9 | 29.5 (25-35) | (31-42) |

### Quality assessment

Based on the results of quality assessment, seven studies had a good quality and three studies had an average quality. The results of the quality assessment are shown in Supplementary 1.

**Supplementary 1:**
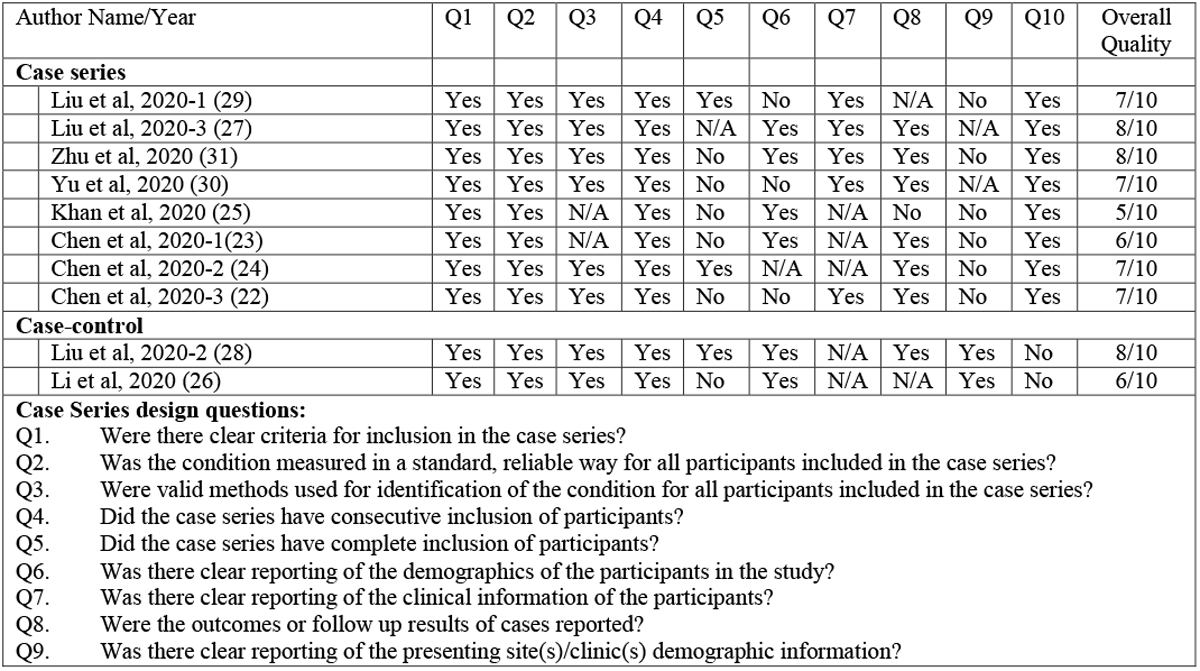
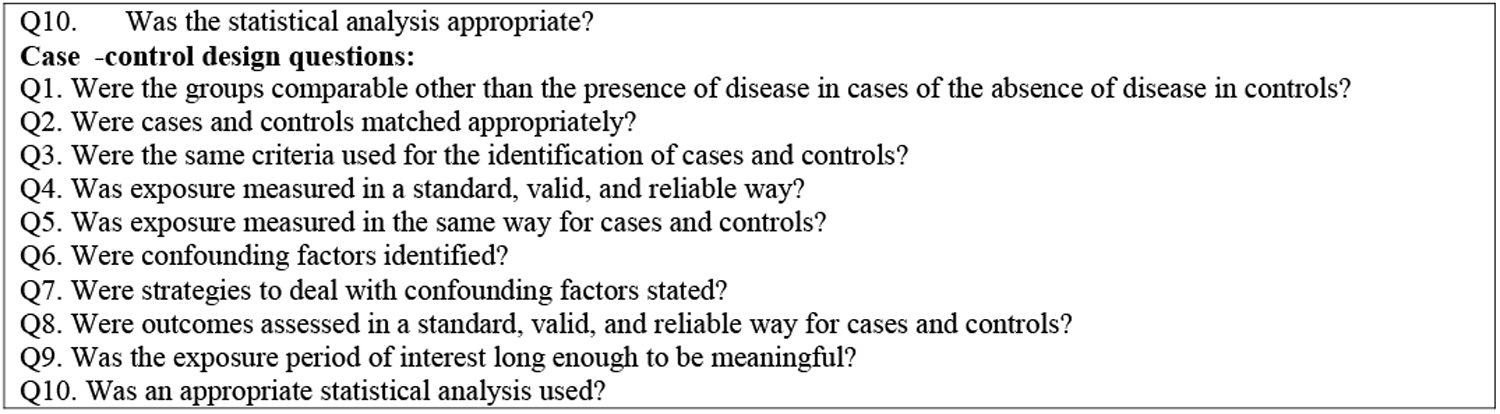
JBI critical appraisal checklist applied for included studies

| Author Name/Year | Q1 | Q2 | Q3 | Q4 | Q5 | Q6 | Q7 | Q8 | Q9 | Q10 | Overall Quality |
| --- | --- | --- | --- | --- | --- | --- | --- | --- | --- | --- | --- |
| **Case series** |  |  |  |  |  |  |  |  |  |  |  |
| Liu et al, 2020-1 (29) | Yes | Yes | Yes | Yes | Yes | No | Yes | N/A | No | Yes | 7/10 |
| Liu et al, 2020-3 (27) | Yes | Yes | Yes | Yes | N/A | Yes | Yes | Yes | N/A | Yes | 8/10 |
| Zhu et al, 2020 (31) | Yes | Yes | Yes | Yes | No | Yes | Yes | Yes | No | Yes | 8/10 |
| Yu et al, 2020 (30) | Yes | Yes | Yes | Yes | No | No | Yes | Yes | N/A | Yes | 7/10 |
| Khan et al, 2020 (25) | Yes | Yes | N/A | Yes | No | Yes | N/A | No | No | Yes | 5/10 |
| Chen et al, 2020-1(23) | Yes | Yes | N/A | Yes | No | Yes | N/A | Yes | No | Yes | 6/10 |
| Chen et al, 2020-2 (24) | Yes | Yes | Yes | Yes | Yes | N/A | N/A | Yes | No | Yes | 7/10 |
| Chen et al, 2020-3 (22) | Yes | Yes | Yes | Yes | No | No | Yes | Yes | No | Yes | 7/10 |
| **Case-control** |  |  |  |  |  |  |  |  |  |  |  |
| Liu et al, 2020-2 (28) | Yes | Yes | Yes | Yes | Yes | Yes | N/A | Yes | Yes | No | 8/10 |
| Li et al, 2020 (26) | Yes | Yes | Yes | Yes | No | Yes | N/A | N/A | Yes | No | 6/10 |
| **Case Series design questions:** |  |  |  |  |  |  |  |  |  |  |  |
| Q1. Were there clear criteria for inclusion in the case series? |  |  |  |  |  |  |  |  |  |  |  |
| Q2. Was the condition measured in a standard, reliable way for all participants included in the case series? |  |  |  |  |  |  |  |  |  |  |  |
| Q3. Were valid methods used for identification of the condition for all participants included in the case series? |  |  |  |  |  |  |  |  |  |  |  |
| Q4. Did the case series have consecutive inclusion of participants? |  |  |  |  |  |  |  |  |  |  |  |
| Q5. Did the case series have complete inclusion of participants? |  |  |  |  |  |  |  |  |  |  |  |
| Q6. Was there clear reporting of the demographics of the participants in the study? |  |  |  |  |  |  |  |  |  |  |  |
| Q7. Was there clear reporting of the clinical information of the participants? |  |  |  |  |  |  |  |  |  |  |  |
| Q8. Were the outcomes or follow up results of cases reported? |  |  |  |  |  |  |  |  |  |  |  |
| Q9. Was there clear reporting of the presenting site(s)/clinic(s) demographic information? |  |  |  |  |  |  |  |  |  |  |  |

Q10. Was the statistical analysis appropriate?

### Heterogeneity

Based on the data analysis, a high level of heterogeneity was not observed in the findings. In some cases with the high heterogeneity, the random effect was used. The results of the heterogeneity of included studies are shown in Table II.

**Table II:** Results of heterogeneity among included studies

| Variable | Sub-groups | # of studies | Event rate (%)<br>95% CI | Q-value | Df (Q) | I<sup>2</sup> | P-value | Selected model |
| --- | --- | --- | --- | --- | --- | --- | --- | --- |
| Signs and symptoms | Cough | 9 | 35.5, (23.1-50.2) | 13.93 | 8 | 42.60 | 0.083 | Random |
|  | Diarrhea | 6 | 7.6 (3.3-16.5) | 2.65 | 5 | 0.0 | 0.754 | Random |
|  | Dyspnea | 9 | 14.6 (9.2-22.3) | 2.55 | 8 | 0.0 | 0.895 | Random |
|  | Fatigue | 3 | 14.9 (7-29.1) | 2.79 | 2 | 28.52 | 0.247 | Random |
|  | Fever | 10 | 66.8 (48.3-81.2) | 26.63 | 9 | 66.2 | 0.002 | Random |
|  | Myalgia | 3 | 24.6 (12.1-43.5) | 0.59 | 2 | 0.0 | 0.744 | Random |
|  | Sore throat | 4 | 11.5(4.8-25.1) | 1.71 | 3 | 0.0 | 0.633 | Random |
|  | Post-partum fever | 4 | 37.1 (18.5-60.6) | 6.01 | 3 | 60.1 | 0.016 | Random |
| Laboratory tests | Lymphopenia | 7 | 49.8 (30.1-69.6) | 17.59 | 6 | 65.90 | 0.007 | Random |
|  | Leukocytosis | 4 | 47.7 (31.6-64.2) | 4.57 | 3 | 34.38 | 0.206 | Random |
|  | Elevated neutrophil ratio | 3 | 83.7 (72.3-91.0) | 0.236 | 2 | 0.0 | 0.889 | Random |
|  | Elevated C-reactive protein | 7 | 57 (43.7- 69.3) | 8.89 | 6 | 32.53 | 0.180 | Random |
|  | Decreased lymphocyte ratio | 3 | 71.4 (16.4-96.9) | 19.47 | 2 | 89.73 | < 0.001 | Random |
| Type of delivery | Cesarean | 10 | 84 (74.0-90.7) | 9.76 | 9 | 7.84 | 0.370 | Random |

## Synthesis of results

### Signs and symptoms

Various signs and symptoms have been reported in studies. Of these, the highest was fever with 66.8 % (95 % CI; 48.3-81.2). Other signs and symptoms based on the highest percentage: post-partum fever (37.1 %, 95 % CI; 18.5-60.6), cough (35.5.9 %, 95 % CI; 23.1-50.2), myalgia (24.6 %, 95 % CI; 12.1-43.5), fatigue (14.9%, 95 % CI; 7-29.1), dyspnea (14.6 %, 95 % CI; 9.2-22.3), sore throat (11.5%, 95 % CI; 4.8-25.1) and diarrhea (7.6 %, 95 % CI; 3.3-16.5) (Figure 2, A-H and Table II).

**Figure 2:**
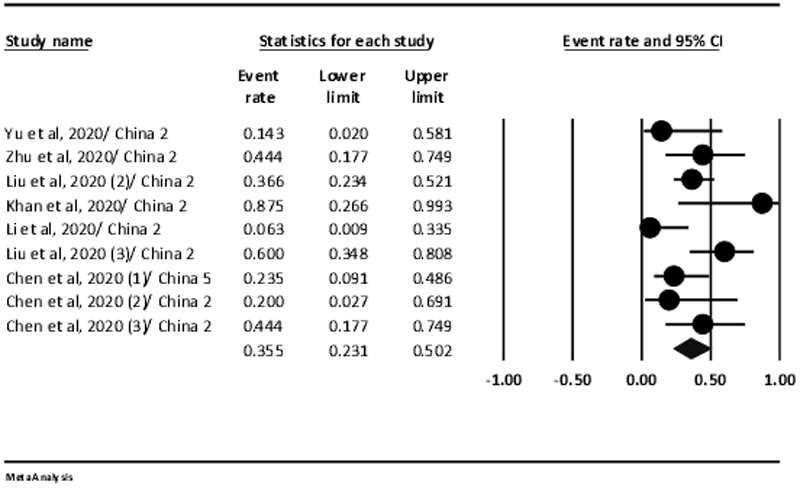
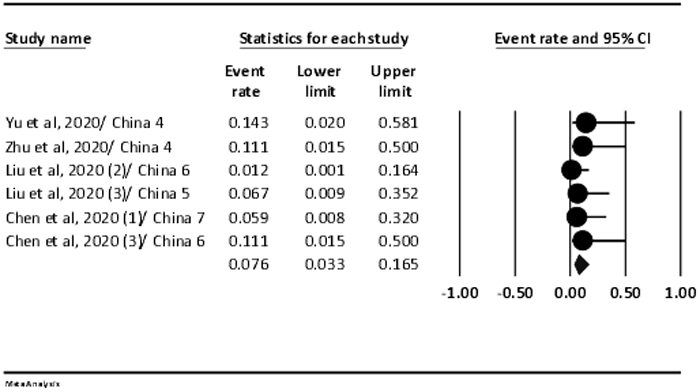
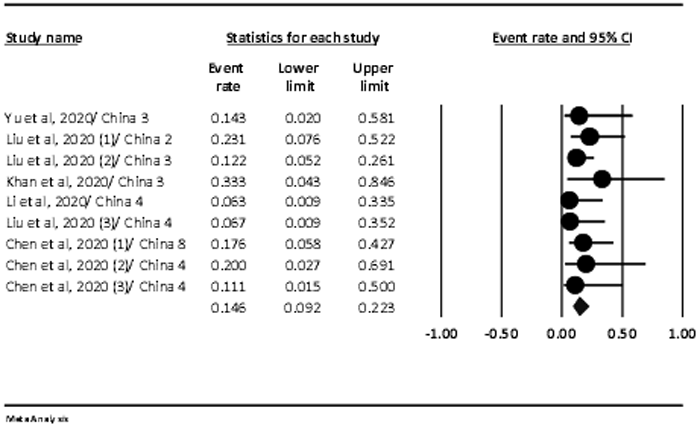
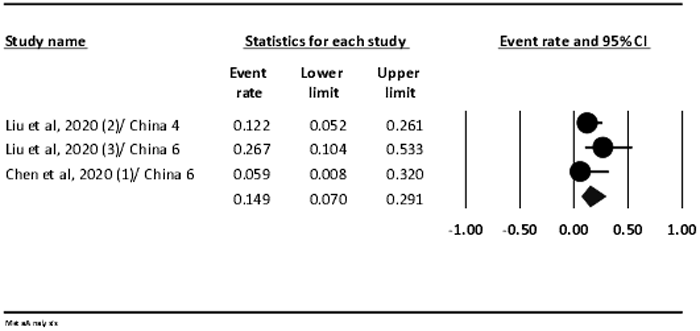
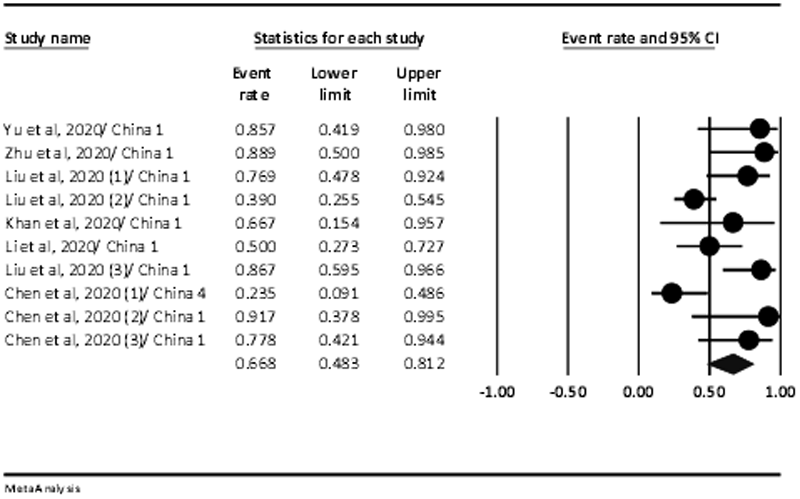
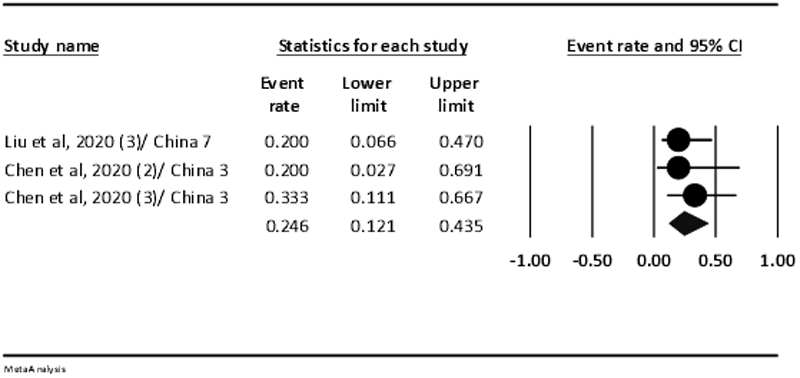
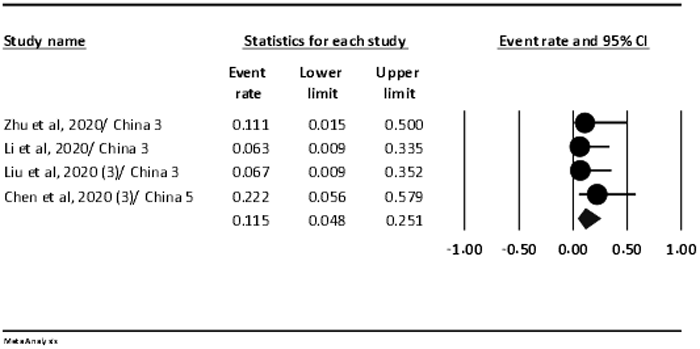
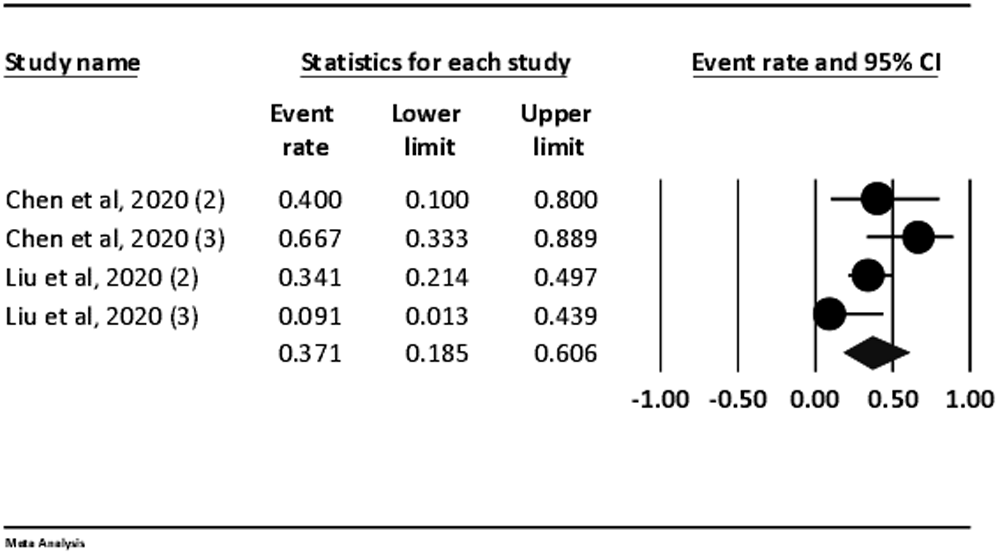
The forest plot presenting event rate and 95% CI for the signs and symptoms in pregnant women with COVID-19; (A) cough, (B) diarrhea, (C) dyspnea, (D) fatigue, (E) fever, (F) myalgia, (G) post-partum fever, and (H) sore throat.

### Laboratory tests

Based on data analysis, lymphopenia with 49.8 % (95 % CI, 30.1-69.6), leukocytosis 47.7 % (95 % CI, 31.6-64.2), elevated neutrophil ratio 83.7 % (95 % CI, 72.3-91.0), elevated C-reactive protein 57 % (95 % CI, 43.7-69.3), and decreased lymphocyte ratio 71.4 % (95 % CI, 16.4-96.9) was observed in studies (Figure 3, A-E and Table II).

**Figure 3:**
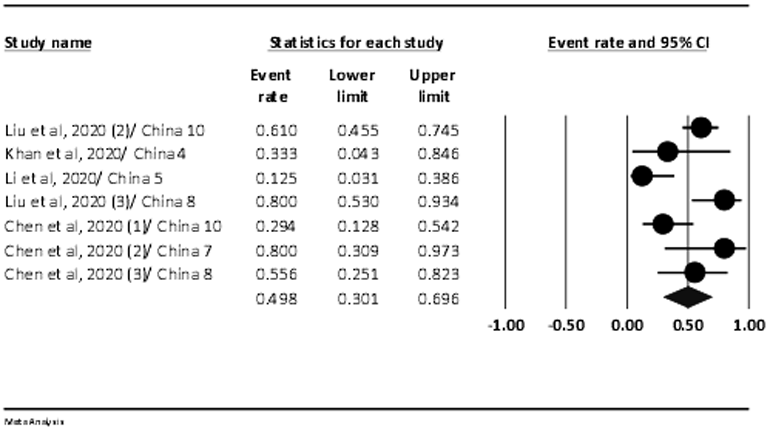
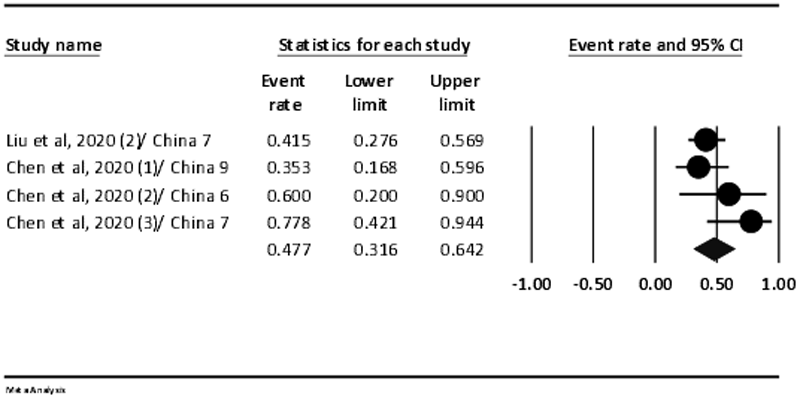
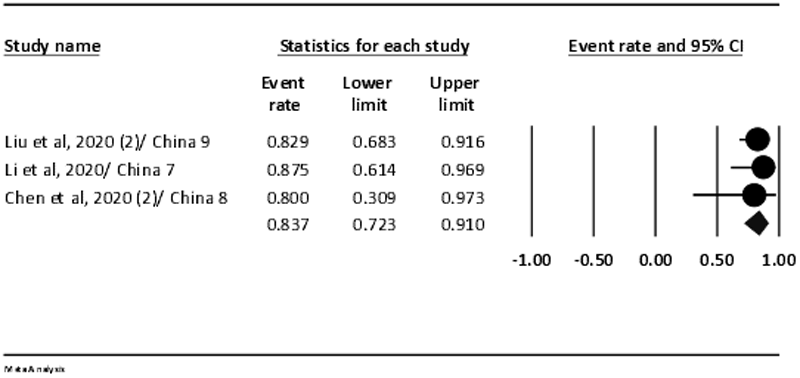
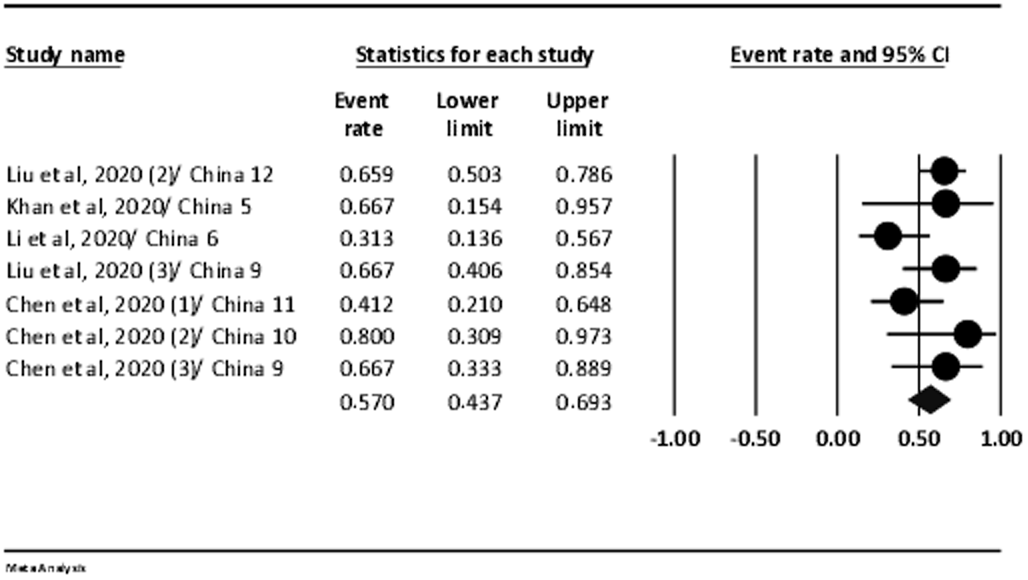
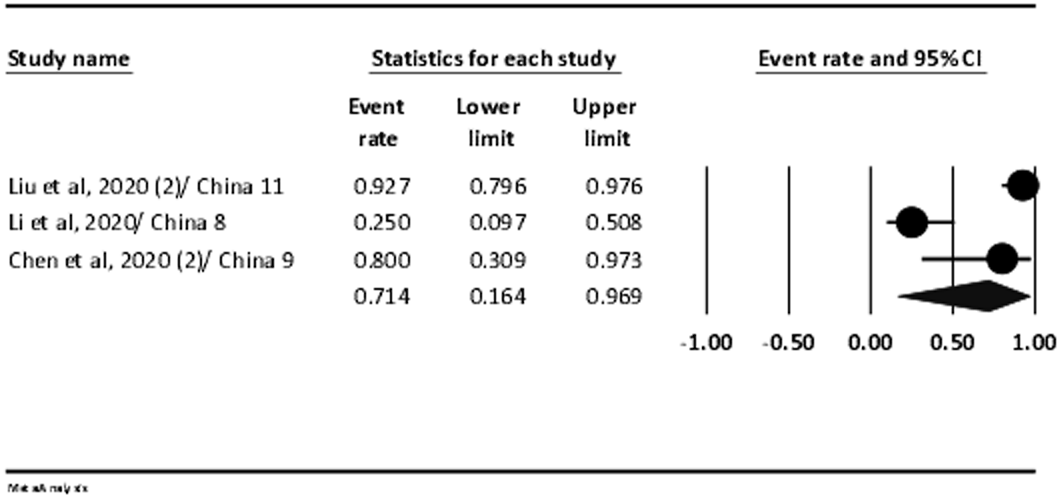
The forest plot presenting event rate and 95% CI for the laboratory tests in pregnant women with COVID-19; (A) lymphopenia, (B) leukocytosis, (C) elevated neutrophil ratio, (D) elevated C-reactive protein, and (E) and decreased lymphocyte ration.

## Type of delivery

According to the results, the rate of cesarean section for delivery in all cases was 84% (95 % CI; 74 - 90.7) (Figure 4 and Table II)

**Figure 4:**
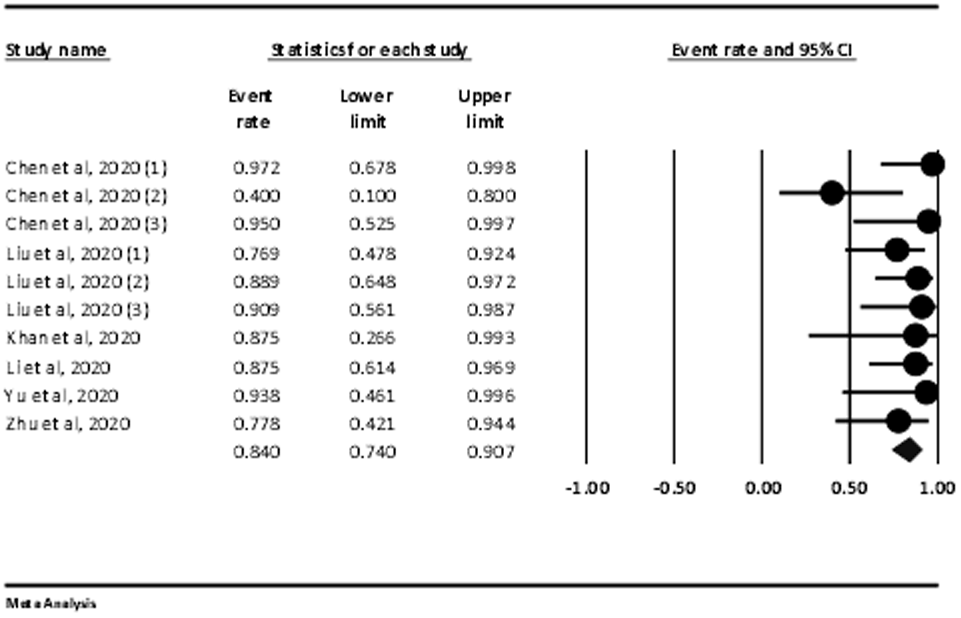
The forest plot presenting event rate and 95% CI for the type of delivery in pregnant women with COVID-19.

### Perinatal outcomes

According to the results, only one case was the newborn of a mother with COVID-19 positive. Also, there was only one death due to DIC. Perinatal outcomes of pregnant women with COVID-19 were shown (Table III).

**Table III.** Perinatal outcomes of pregnant women with COVID-19

| Author, Year (reference number) | Total sample size | Covid-19 Positive | Low Birth weight | Premature | Complication | Outcome (Died) |
| --- | --- | --- | --- | --- | --- | --- |
| Liu et al, 2020-1(29) | 13 | 0 | NR | 6 | 0 | 0 |
| Liu et al, 2020-2 (28) | NR | NR | NR | NR | NR | NR |
| Liu et al, 2020-3 (27) | 2 | 0 | NR | 0 | 0 | 0 |
| Zhu et al, 2020 (31) | 10 | 0 | 7 | 5 | Multiple organ failure and DIC (1) | 1 |
| Yu et al, 2020 (30) | 7 | 1 | 0 | NR | 0 | 0 |
| Khan et al, 2020 (25) | 3 | 0 | 0 | 1 | 0 | 0 |
| Li et al, 2020 (26) | 17 | 0 | 3 | 4 | Intrauterine fetal distress (2) | 0 |
| Chen et al, 2020-1 (23) | 17 | 0 | 0 | 0 | 0 | 0 |
| Chen et al, 2020-2 (24) | 5 | 0 | 0 | 0 | 0 | 0 |
| Chen et al, 2020-3 (22) | 9 | 0 | 2 | 4 | 0 | 0 |

### Results of meta-regression

According to the findings, the only factor that could be examined in this section was the mean age of pregnant women. Data analysis showed that older pregnant women significantly have a higher fever rate (Coefficient=0.477, P=0.033). For the type of delivery, the higher average age of pregnant women significantly associated with a higher rate in cesarean delivery (Coefficient=0.433, P=0.016). The results of the relationship between other factors and the average age of pregnant women are shown in Table IV(Supplementary 2).

**Table IV:** Result of meta-regression

| Variable | Sub-groups | Mean of age |  |  |
| --- | --- | --- | --- | --- |
|  |  | Coefficient | S.E | P-value |
| Signs and symptoms | Cough | 0.192 | 0.207 | 0.354 |
|  | Diarrhea | 0.103 | 0.392 | 0.792 |
|  | Dyspnea | -0.110 | 0.236 | 0.640 |
|  | Fatigue | 0.676 | 0.405 | 0.095 |
|  | Fever | 0.477 | 0.223 | 0.033 |
|  | Myalgia | 0.0 | 0.322 | 0.999 |
|  | Sore throat | -0.117 | 0.520 | 0.821 |
|  | Post-partum fever | -0.001 | 0.192 | 0.995 |
| Laboratory tests | Lymphopenia | 0.309 | 0.252 | 0.220 |
|  | Leukocytosis | 0.092 | 0.324 | 0.775 |
|  | Elevated neutrophil ratio | 0.141 | 0.443 | 0.749 |
|  | Elevated C-reactive protein | 0.057 | 0.231 | 0.802 |
|  | Decreased lymphocyte ratio | -0.649 | 0.431 | 0.131 |
| Type of delivery | Cesarean | 0.433 | 0.180 | 0.016 |

**Supplement 2:**
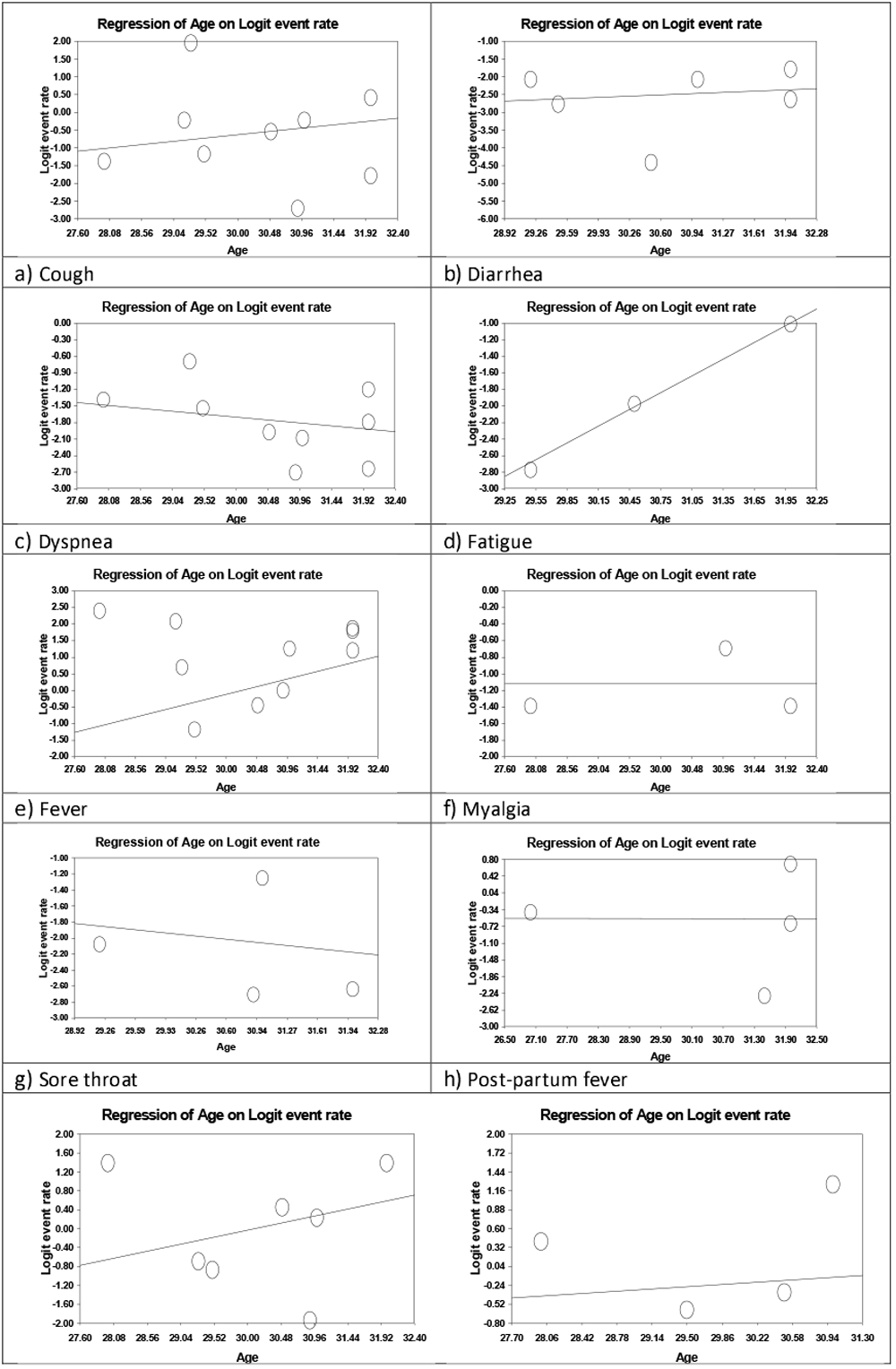
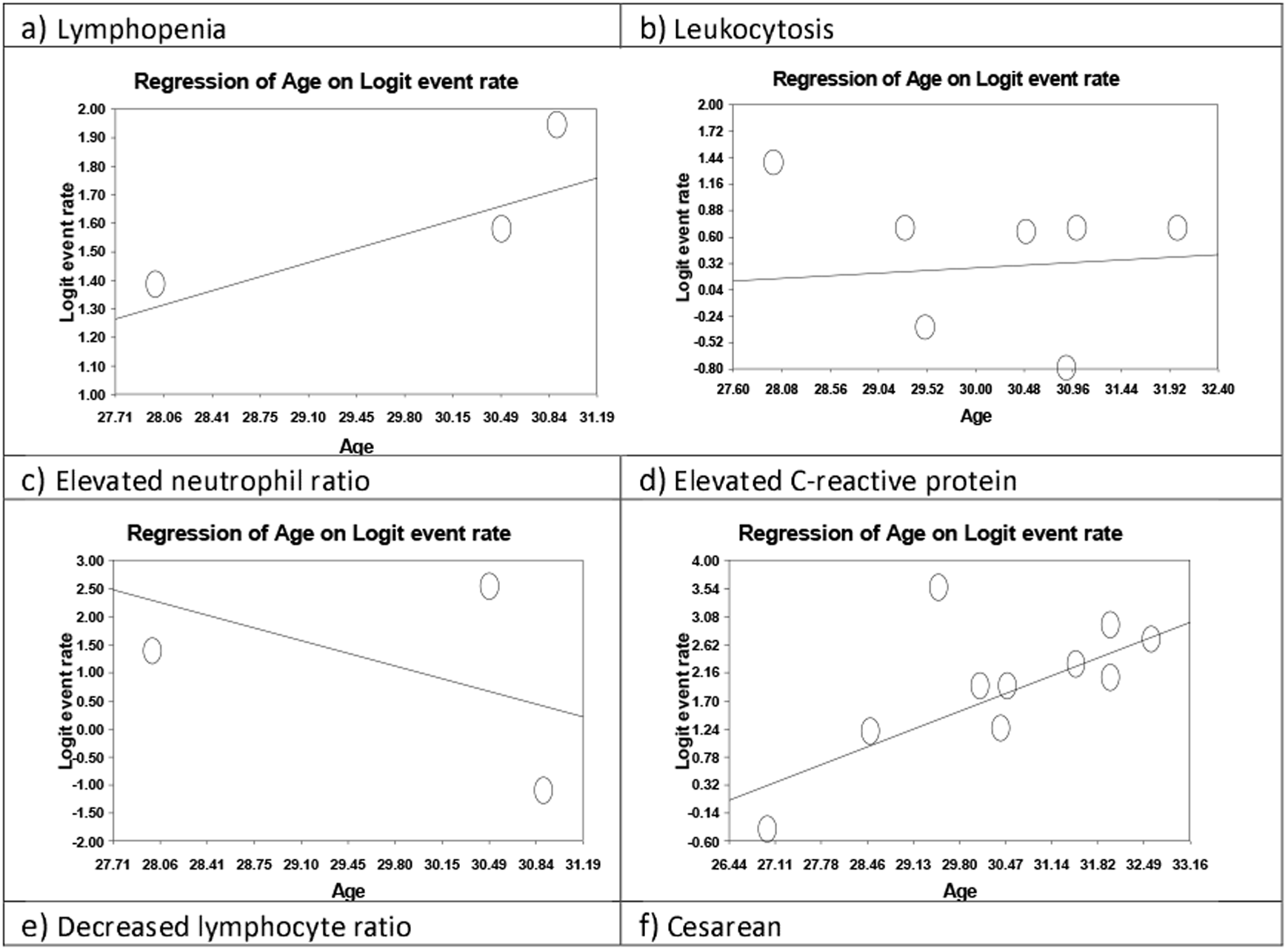
Result of meta-regression

**Supplementary 3:**
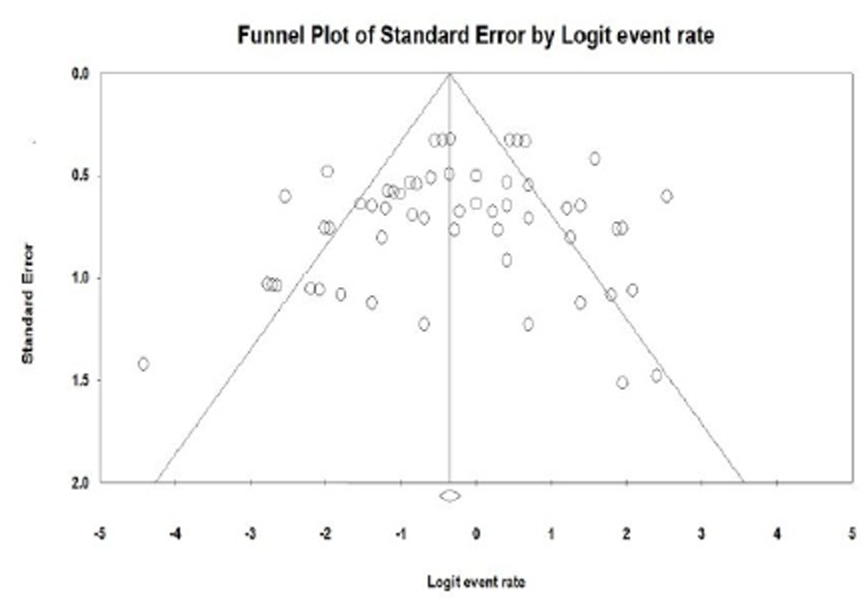
Funnel plot for fever

### Publication bias

Visual inspection of funnel plot and Egger’s tests did not indicate evidence of publication bias (P=0.127) (Supplementary 3).

## Discussion

A total of 10 articles were reviewed in this study, which finally analyzed 135 pregnant women, all of whom were in the third trimester of pregnancy (22-31). These summary findings help healthcare workers better manage pregnant women with COVID-19, which could potentially reduce the side effects for women as well as their newborns.

Common clinical manifestations of pregnant women with COVID-19 include fever and cough, and less common symptoms were sore throat and diarrhea. Postpartum fever is also more common in women after childbirth. However, the rate of fever in our study was lower than that of Guan and colleagues, who studied the symptoms of non-pregnant coronary artery disease and reported a rate of fever of 87.9 percent. But in this study, as in our study, diarrhea was the least common (32).

In terms of laboratory demonstrations, Elevated neutrophil ratio and Decreased lymphocyte ratio have been common. On the other hand, the prevalence of CRP Elevated in our study was 57 %. However, in Zhang’s and co-authors study, this prevalence in a group of people with non-severe and severe patients was 88.9 and 96.4%, respectively (33). This indicates a more pronounced inflammation in patients with more severe conditions and given that pregnant women in this study were not in severe disease conditions, a lower percentage of increased CRP prevalence is justified. On the other hand, in Rodriguez-Morales co-workers study, the increased CRP prevalence was 58.3, which is similar to our study (8). These differences in numbers can be described due to the severity of the disease, and on the other hand, a more comprehensive examination is needed.

Lymphopenia and Leukocytosis were less common in our study. However, in the study of Zhang and colleagues and Wang et al, which was performed on patients with corona (normal population), lymphopenia was the most common laboratory symptom and was 75.4 and 70.3%, respectively (33, 34). However, it should be noted that these numbers are a decrease in absolute lymphocyte count.

In our study, the majority of pregnancies have happened cesarean section, which is much higher than the World Health Organization’s recommendation for vaginal route delivery (35), which can be determined by a gynecologist to prevent maternal respiratory distress during pregnancy.

In our current study, which examined 135 pregnant women with COVID-19 pneumonia, none of the patients with severe or dead pneumonia were infected with COVID-19 infection. Although SARS-CoV-2 has a common sequence with SARS of up to 85%, we need to be aware of the possibility that the course of the disease and the prognosis of this disease can follow the same SARS process in pregnant women (36-37).

Our study has some limitations. First, all patients registered in these included articles were in the third trimester of pregnancy, and the effect of the virus infection on the fetus in the first or second trimester was unknown. Second, due to the short duration of the outbreak, the long-term consequences of the disease on infants have not been possible and more studies are needed. Third, the low number of samples of articles included is another limitation of the work. Fourth, all included studies were from china.

In conclusion, pregnant women with COVID-19 pneumonia had diverse symptoms; however, fever and cough are the main clinical symptoms of these women. Although one infant was born with COVID-19 in the included studies, there was little evidence that COVID-19 was transmitted from mother to infant in late pregnancy. Therefore, the study of long-term outcomes on mother and child, as well as the vertical transfer of mother to child in second-trimester pregnancies and the first months after delivery, requires further studies.

## Data Availability

there is not data available

